# Differences in innate Intracellular viral suppression competencies may explain variations in morbidity and mortality from SARS-CoV-2 infection

**DOI:** 10.1101/2020.09.13.20193524

**Authors:** Shaibu Oricha Bello, Ehimario Igumbor, Yusuf Yahaya Deeni, Chinwe Lucia Ochu, Popoola Mustapha Ayodele

**Affiliations:** Department of Pharmacology & Therapeutics, College of Health Sciences, Usmanu Danfodiyo University, PMB 2346, Sokoto, Nigeria; Nigeria COVID-19 Research Consortium, Nigeria Centre for Disease Control, Abuja-Nigeria; Nigeria Center for Disease Control, Abuja, Nigeria; School of Public Health, University of the Western Cape, Cape Town, South Africa; Department of Microbiology & Biotechnology, Federal University Dutse, PMB 7156, Jigawa State, Nigeria; Center for Environmental and Public Health Research and Development (CEPHARD), Kano, Kano State, Nigeria; Research & Development Matters, Tertiary Education Trust Fund, Abuja

## Abstract

SARS-CoV-2 infection and COVID-19 ravage the world with wide variations in morbidity and mortality that have remained largely unexplained, even by mutations in protein coding regions. In this study, we analyzed available complete SARS-CoV-2 sequences using the CpG index as a signature of Zinc finger antiviral protein (ZAP) activity to examine population variations in innate intracellular antiviral competencies. The result suggests that differential ZAP activity may be a major determinant of the outcome of SARS-CoV-2 infection. SARS-CoV-2 sequences from Africa, Asia, and pools of asymptomatic patients had I_CpG signature evidence of high ZAP activity, while SARS-CoV-2 sequences from North America and Intensive Care Unit or Deceased patients had I_CpG signature of low ZAP activity. ZAP activity is linked to the interferon system. Low ZAP activity may be part of the explanation for the increased morbidity of SARS-CoV-2 in the elderly and with comorbidities like diabetes, obesity, and hypertension. It may also provide some insight into the discrepancies between invitro anti-SARS-CoV-2 activities of candidate therapies and performance in clinical trials. Furthermore, our results suggest that asymptomatic patients may paradoxically shed a more dangerous virus.

## Introduction

New Severe Acute Respiratory Syndrome Coronavirus (SARS-CoV-2) infection was first reported in Wuhan China in late December 2019 but has rapidly spread across the world bringing in its wake a disease, COVID-19, that has challenged the best health systems[1][2]. While the world of science and medicine have responded with rapid research into this newest microbial challenge and has indeed garnered some insight into SARS-CoV-2 and COVID-19, large uncertainties remain. Overall, SARS-CoV-2 infection remains largely asymptomatic (80%) in most persons and causes mild, moderate(15%), or severe illness(5%) in others with no clear explanation for the spectrum of illness[3]. Furthermore, the predicted decimation by COVID-19 of populations with highly underdeveloped and unprepared health systems appears widely off-track[4][5][6]. The highest cases of SARS-CoV-2 infections, most severe COVID-19 and highest crude case fatality rates/ mortalities are in Europe and North America[7][8]. Overall, COVID-19 mortality varies regionally between 0.8% to 14%[9]. Africa appears to have been largely spared of its worst-case scenarios[10], a situation currently described as mysterious[11] in search of a better explanation. It has been suggested that the relatively high rates of morbidities like diabetes and hypertension (both known to worsen COVID-19) in Europe and the Americas on the one hand, and the high rate of cross-protecting vaccinations (like BCG) and cross-protecting immunity from rampant endemic infections in Africa on the other hand, may have contributed to the differing outcomes of SARS-CoV-2 infections and COVID-19 in these geographical regions[12]. Emerging serological studies suggest that indeed, high rates of SARS-CoV-2 infection are ongoing in Africa with some studies suggesting a background rate of 9.3% of some African population seropositive[13]. Contrary to prediction that incidence intensity should predict COVID-19 mortality, as suggested from studies[14], the fatality was 8 times less than predicted among a group of seropositive health care workers in Africa[15]. Together, these findings suggest that, although SARS-CoV-2 is indeed spreading in Africa as predicted, certain post-infection events mitigate COVID-19 while at the same time allowing continuing transmission. One possible explanation is post-infection cellular attenuation of SARS-CoV-2 by natural intracellular antiviral mechanisms. One such cellular antiviral mechanism, whose activity signature may be traceable, is Zinc Finger Antiviral Protein (ZAP)[16].

After entering cells, viruses must overcome the intracellular antiviral machinery to establish a productive infection. In response to pathogen detection, host cells produce type 1 Interferons (IFNs) which signals downstream through JAK/STAT to induce numerous IFNs-Stimulated genes (ISGs) that inhibit various steps in the life cycle of the virus[17]. ZAP is one such ISG that is particularly effective against Beta coronaviruses[16]. ZAP, also called PARP13 (poly (ADP-ribose) polymerase 13), is alternatively spliced and produced as the long form (ZAPL) and short form (ZAPS)[17]. ZAPS is upregulated more than ZAPL by RNA virus infections and has been shown to restrict SARS-CoV-2 in human cell-lines known to be targets of the virus[16]. ZAP crucially inhibits translation of incoming viral RNA (an important action against positive sense RNA viruses) and synergizes with other ISGs to upregulates RIG-1 IFNs-Beta production[17]. Precisely, ZAP uses CpG dinucleotide (5’-Cytosine-phosphate-Guanosine-3’) on viral sequences as targets for recognition, binding, and then recruits the exosome complex to degrade viral genome[18][19]. Viruses with high CpG content are therefore more susceptible to ZAP restriction, while viruses with low CpG content are less susceptible[19]. It has been shown that SARS-CoV-2 may have been able to cross to human host by acquiring CpG deficiency[20] and thereby mimic the CpG deficiency of human genome[21]. The null expectation of CpG index (I_CpG) is 1[20][22], and less than or more than 1 are considered deficiency and overrepresentation [22]respectively; though it has also been suggested that I_CpG of 0.78 should be taken as significant deficiencies and I_CpG 1.22 should be taken as significant overrepresentation[23]. In this schema, <0.5 is considered extremely low and >1.5 extremely high[23]. It has been stated that “SARS-CoV-2 has the most extreme CpG deficiency of all known Beta coronavirus genomes”[20]. The I_CpG of the reference Wuhan SARS-CoV-2 is 0.4077 and is ‘extremely deficient’ according to the criteria previously stated[23]. It has been shown that ZAP knockdown in cells increases SARS-CoV-2 production by 3 folds and that this increase can be as high as 7.7 folds in the presence of Interferons[24]. Given that ZAP exhibits tissue-specific expressions[20], the resulting difference in intracellular antiviral activity can be predicted to leave CpG signatures on SARS-CoV-2 and that I_CpG variations will reflect the spectrum of tissues infected[20]. Such CpG signatures may therefore be used to identify viral tissue-switching events[20].

It may be predicted that given that SARS-CoV-2 infection has occurred, several ZAP induced CpG restriction scenarios may occur. In host communities with globally highly effective ZAP, COVID-19 will be milder, and this will also reveal as less variable SARS-CoV-2 I_CpG evolution within the community. On the contrary, in host communities with less effective ZAP suppression, COVID-19 will be more severe, and this will reveal as more variable SARS-CoV-2 I_CpG spectrum. Furthermore, within a host, a highly variable ZAP expression spectrum may result in more widespread tissue affectation with only host tissues with the higher order ZAP expression being spared.

APOBEC3G (apolipoprotein B (APOB) mRNA-editing, catalytic polypeptide), is another natural antiviral factor that targets CpG to restrict RNA viruses[25]. APOBEC3G deaminates C to U and may contribute to CpG deficiency[26]. Such C to U conversion is deleterious to the viral fitness if the deaminated site is in a functional region but may reduce viral susceptibility to CpG guided antiviral activity of ZAP[20]. Though unlike ZAP, APOBEC3G is not highly specific for cytosine[20], high activity of APOBEC3G on SARS-CoV-2 may drive UpG overexpression (i.e. I_UpG>1)[27].

In totality, if intracellular antiviral activity related to CpG or UpG restrictions or overexpression respectively are significant events in COVID-19, then such signatures will be expected in SARS-CoV-2 sequences in sample populations. This study was therefore conducted to determine the I_CpG and I_UpG signature patterns in available SARS-COV-2 sequences and identify any relationship with disease severity.

## Methods

### Data-acquisition and Data Treatment

SARS-CoV-2 sequences were obtained from GenBank[28]. GenBank was chosen as the primary data source because it has been assessed as exceptionally reliable[29]. All complete SARS-CoV-2 sequences from human host on GenBank on 1st August 2020, were downloaded as a single file without geographical restrictions and then downloaded again with geographical restrictions as separate files per region. The regions were taken as allocated on GenBank-namely, Africa, Asia, Europe, Oceania, North America (NAM), and South America (SAM). Complete SARS-CoV-2 sequences that contained patient metadata was downloaded from GISAID[30]. Such data was not available on GenBank. On both platforms, SARS-CoV-2 sequences were downloaded as FASTA format, but on the GISAID platform, patient metadata was downloaded in a separate tab separated values(tsv) file. Sequences from GISAID were then matched to patient metadata on Microsoft excel sheet using ‘sequences name’ and ‘virus name’ respectively. Matched data were visually cross-checked for mismatch. Seventeen mismatches were identified and deleted before analysis. The treated GenBank data and GISAID patients matched data are available as supplementary files 1 and 2 respectively on this article. The reference SARS-CoV-2 viral sequence was NC_045512.2 SARS-CoV-2 Wuhan genome. COVID-19 crude case fatality rate for different geographical regions as of 1^st^ August 2020 was obtained from ‘Our world in data’ (OWID). Data from this site has been rated left-center biased but highly factual [31], [32], and has been cited in various high impact medical journals[33], [34]. Again, the geographical regions were defined as Africa, Asia, Europe, Oceania, North America (NAM), and South America (SAM). On the OWID website, the crude case fatality at any point in time can be read-off by placing the cursor on the desired date.

### Data analysis & Statistics

Sequence analysis and calculations were implemented on DAMBE7 software[35]. NCSS2020 statistical software, Analyze-IT-excel add-on, and JASP 0.13.1 were used for statistical analysis. In a study of the geographic and genomic distribution of SARS-CoV-2 mutation, a minimum of 50 sequences was considered as adequate for analysis [36]but we pre-specified an expected variability of 5% in the I_CpG and at a power of 80% and alpha of 5%, which gave a sample size of 73. Therefore, only regions with above 73 deposited SARS-CoV-2 sequences were included in the analysis(this excluded SAM, that had only 18 deposited complete sequences from human host as of 1^st^ August 2020). All complete sequences were used in the analysis but for sensitivity, the analysis was repeated with sequences with less than NNNs. We included only patients’ status labeled ‘Asymptomatic’(Asy), ‘Intensive Care Unit’ (ICU) or ‘Deceased’(D) in the analysis because these terms were considered non-ambiguous. ICU and Deceased were merged as one group (ICU/Deceased) for analysis because these two were considered as one extreme of COVID-19 severity and that the difference is likely linked to therapy rather than viral factor. Also, ICU and Deceased status have been merged in previous categorical scaling of COVID-19 [37]. Patients’ status like ‘Alive’, ‘Hospitalized’ and ‘Discharged’ were available on the metadata file, but such patient and corresponding sequenced were excluded from analysis because these terms were not classifiable as regards COVID-19 severity. Only extreme SARS-CoV-2 infection status (Asy vs ICU/Deceased) was compared because these were more clearly defined and were more likely to reveal differences in dinucleotide status if these exist. Another parameter of interest was viral Clade.

I_CpG was calculated as 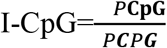. I_UpG was similarly calculated. The I_CpG and genomic GC proportion(GC%) was initially suggested to be a mathematical artifact[38] but this has been refuted[39]. Furthermore, it has been shown that differential CpG altering events alter the correlation between I_CPG and GC content in a genome[39]. We expected that both intracellular antiviral activities and random mutations may alter I_CpG. We hypothesized that highly variable intracellular ZAP antiviral activity in a host population will lead SARS-CoV-2 ‘quasi-species’ with altered I-CpG vs GC% correlation. Absolute GC% was therefore calculated, as well as the correlation between I_CpG and GC% as another sign of ZAP activity. The population (total sample) rate of change of I_CpG with GC% was determined by linear regression and taken as the background rate.

Categorical variables were analyzed using non-parametric statistics. All continuous variables failed normality tests and were therefore analyzed using non-parametric statistics. However, because population (and not sample) distribution is the superior consideration for choice of statistic and this is unknown for SARS-CoV-2, sensitivity analysis was performed by running parametric statistics for comparison. Where the outcomes of the analysis were contradictory, the result of the non-parametric analysis was used for decision. Spearman correlation was used to quantify the relationship between I_CpG and GC Content. Kruskal-Wallis test was used to compare dinucleotide Indexes between geographical regions and Mann-Whitney test was used to compare dinucleotide Indexes between Asy and ICU/Deceased. Spearman correlation was used to quantify the relationship between crude case fatality rate and dinucleotide indexes; for this comparison summary nucleotide indexes per geographical region were used. Considering that by nature, both the crude case fatality rate of SARS-CoV-2 infection and dinucleotide indexes should evolve with time, we consider such a time point comparison plausible. Mann-Whitney was used to test for a significant difference between the age of ‘Asy’ and ‘ICU/Deceased’ while Chi-Square was used to test for a significant difference in SARS-COV-2 dinucleotide distribution between genders. These two (gender and age) were considered possible confounders because they are known to influence SARS-CoV-2 and COVID-19 morbidity and disease severity respectively[12], though ZAP expression may not vary with age or gender[40]. The slope of change of I_CpG with GC% was determined using linear regression. Contingency tables were developed to compare viral Clade over-representation in any patients’ status and type III Chi-Square was used to assess significant differences. All clades were necessarily those used by GISAID (S, L, V, G, GH, GR)[41]. Clade O was also included in the patients’ metadata though this clade is generic and less clearly defined[42].

It has been shown that adults greater than 65 years account for 80% of COVID-19 hospitalizations and have a 23-fold risk of death compared to those less than 65 years old[43].

Although various age classification methods exist, for this study we used the consensus definitions of both Segen’s Medical Dictionary[44] and McGraw-Hill Concise Dictionary of Modern medicine[45] to define middle age as 45-65 years. We, therefore, defined 2 additional age classes: Young <45 years and elderly >65 years. The level of statistical significance was p<0.05.

Overall, three CpG based ZAP signatures were, therefore, used for comparisons between SARS-CoV-2 groups of sequences: (i)Group’s correlation of I_CpG with GC%, (ii) Group’s rate of change of I_CpG for a given GC%, and (iii) Group’s median I_CpG and within-group variability. Where the data distribution was not amenable to linear regressions, the rate of change of I_CpG per GC content was considered inestimable. Linear regressions were considered suitable for comparison of slope differences when the R^2^ of each slope was higher than 0.3 and the difference between R^2^ did not transcend R^2^ quality classification[46],[47]. As sensitivity analysis to account for the variation of I_CpG with GC content, the I_CpG-GC ratio was determined and compared between groups. As regards UpG, only the group’s median I_UpG and within-group variability were used for comparisons. Group’s central dinucleotides values (e.g. mean, median) were considered poorer comparators for trend because they would be biased by the value of the index case of SARS-CoV-2 infection in a country or region.

## Ethics

This study used anonymized, non-personal, and publicly available data sets. All necessary authorization and permissions were obtained and reporting standards including due acknowledgments have been complied with.

## Results

8693 complete SARS-CoV-2 sequences from GenBank, representing 6729(NAM), 934(Asia),568(Oceania), 360(Europe) and 102 (Africa), and 1 reference SARS-CoV-2 sequence were used for analysis. A total of 349 sequences with patients’ metadata from GISAID were available for analysis but the gender of only 286 patients was available: 172 Male and 114 Female.

### Comparison of variations in correlations of I_CpG and GC content of SARS-CoV-2 with geographical regions

In the background population sequences of SARS-CoV-2, I_CpG correlates with GC% (rs=0.359, 95% CI=0.340-0.378) (Figure 1) but this correlation varies significantly (p<0.05) between geographical regions: Africa (rs=0.357, 95% CI=0.169-0.520), Asia(rs=0.029, 95% CI=-0.037-0.095), Europe(rs=0.107, 95% CI=0.00-0.211), Oceania(rs=0.331, 95% CI=0.254-0.405) and North America (NAM) (rs=0.413, 95% CI=0.392-0.433) (Figure 1).

**Figure 1.**
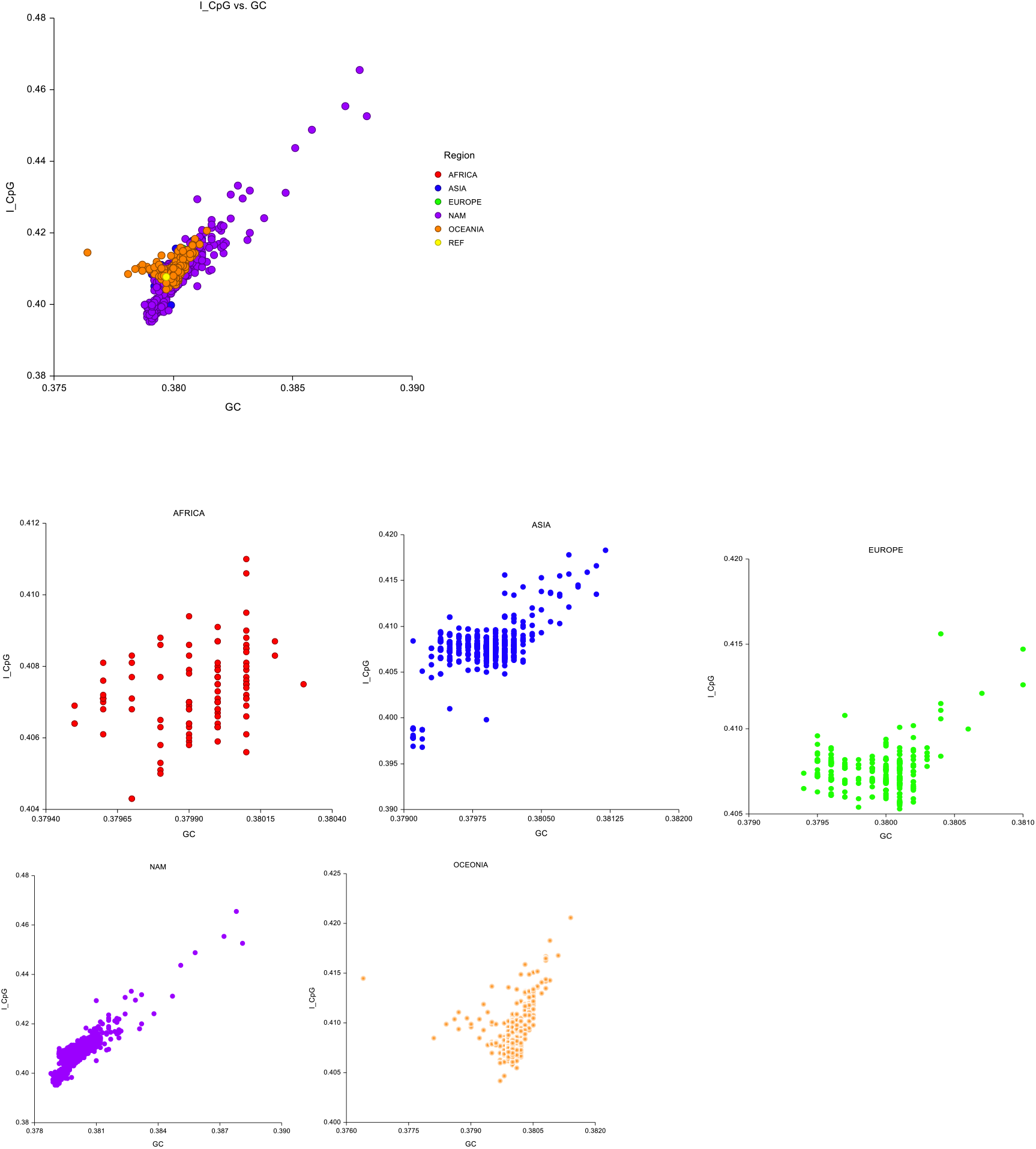
SARS_COV2 I_CpG Vs GC distribution in the study sample. Compared to the Wuhan Virus Reference sequence (in yellow), I_CpG of SARS-CoV-2 has varied widely(Ref=0.4077, Sample range: 0.3952-0.4655, Median 0.4073) and this I_CpG variation is more in NAM and Oceania regions. In the combined sequences, a neat linear regression can be drawn, giving a rate of change of I_CpG for a given value of GC as 7.3 (I_CpG=7.30GC-2.37, R^2^=0.70). This is taken as the population rate.

### Comparison of variations of SARS-CoV-2 dinucleotides frequencies within geographical regions

Compared to the I_CpG of the Wuhan Virus Reference sequence (0.4077), I_CpG of SARS-CoV-2 varied widely (Range: 0.3952-0.4655, Median 0.4073). The I_CpG variation significantly differs between geographical regions (P<0.0001) (Table1a) (Figure 2). Also, Africa has the lowest within region I_CpG deficiencies variations while Asia, NAM, and Oceania had significantly higher variations (p<0.05) (Figure 2). I_UpG was overexpressed in SARS-CoV-2 and significantly differentially overexpressed between geographical regions (P<0.0001) (Table 1) (Figure 2). Africa has the lowest within region I_UpG overexpression variations while NAM and Oceania had significantly higher variations(p<0.05) (Figure 2). Europe has lower I_CpG and I_UPG variations (Figure 2).

**Table 1:** Variation of SARS-COV-2 dinucleotide frequencies with geographical region N=8693

| **I_CpG**<br>**by Region** | **N** | **Minimum** | **1st Quartile** | **Median** | **3rd Quartile** | **Maximum** |
| --- | --- | --- | --- | --- | --- | --- |
| AFRICA | 102 | 0.4043 | 0.40659 | 0.40710 | 0.40800 | 0.4110 |
| ASIA | 934 | 0.3968 | 0.40720 | 0.40760 | 0.40810 | 0.4183 |
| EUROPE | 360 | 0.4053 | 0.40700 | 0.40755 | 0.40800 | 0.4156 |
| NAM | 6729 | 0.3952 | 0.40660 | 0.40720 | 0.40770 | 0.4655 |
| OCEANIA | 568 | 0.4042 | 0.40740 | 0.40800 | 0.40890 | 0.4206 |
| P<0.0001 (Kruskal-Wallis test) |  |  |  |  |  |  |
| **I_UpG**<br>**by Region** | **N** | **Minimum** | **1st Quartile** | **Median** | **3rd Quartile** | **Maximum** |
| AFRICA | 102 | 1.3740 | 1.37530 | 1.37600 | 1.37650 | 1.3779 |
| ASIA | 934 | 1.3679 | 1.37500 | 1.37560 | 1.37610 | 1.3806 |
| EUROPE | 360 | 1.3711 | 1.37570 | 1.37620 | 1.37680 | 1.3779 |
| NAM | 6729 | 1.3583 | 1.37500 | 1.37550 | 1.37620 | 1.3871 |
| OCEANIA | 568 | 1.3709 | 1.37490 | 1.37560 | 1.37620 | 1.3973 |
| P<0.0001 (Kruskal-Wallis test) |  |  |  |  |  |  |

**Figure 2.**
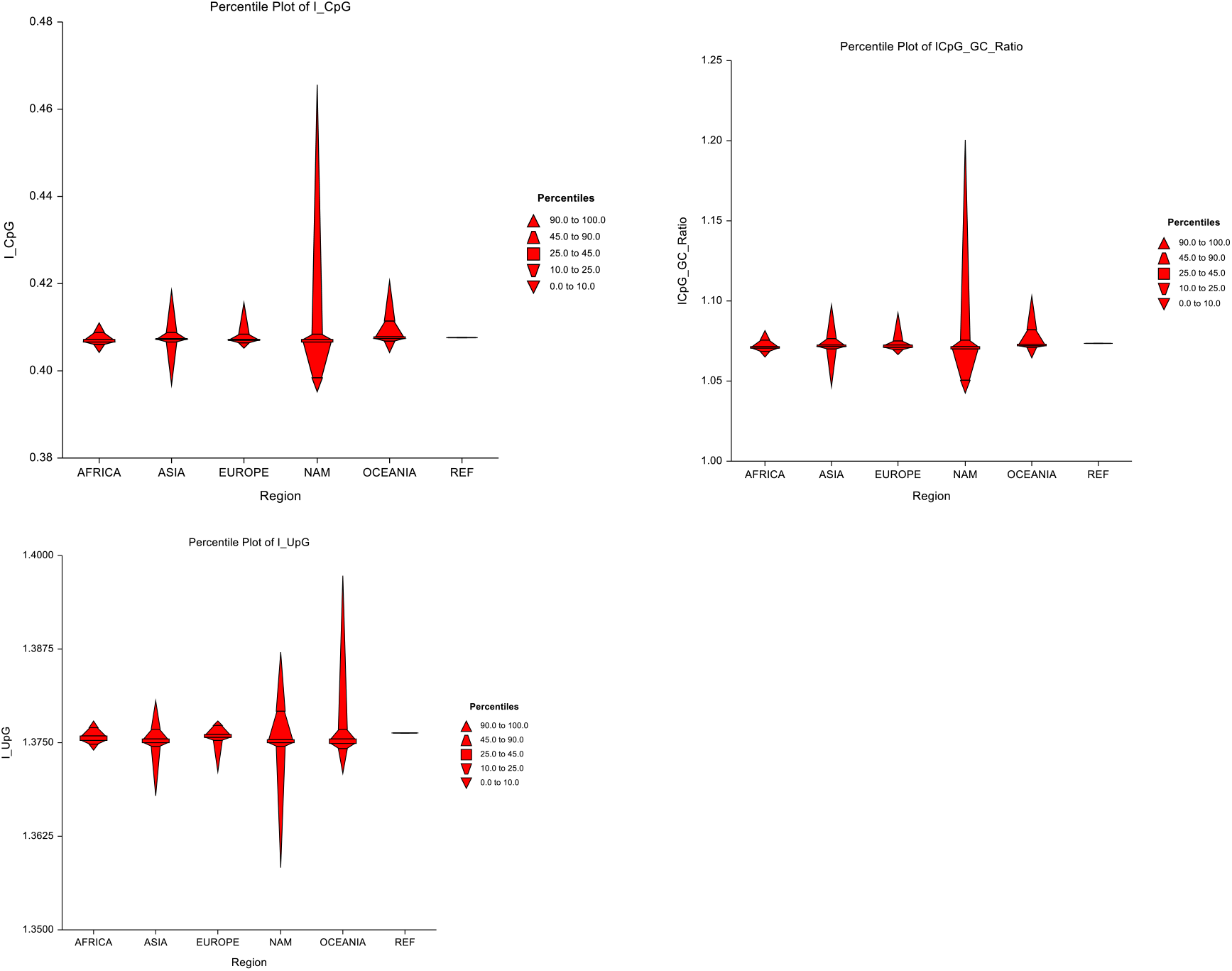
Percentile plot of dinucleotides per geographical region. SARS-CoV-2 sequences from Africa show the lowest I_CpG variability suggestion highly restrictive (competent) ZAP activity.

### Comparisons of variations of SARS-COV-2 dinucleotides frequencies and crude case fatality rates of geographical regions

As of 1^st^ August 2020, 4 crude case fatality rates were available for 5 geographical regions: 1.2(Oceania), 2.1 (Africa and Asia), 4 (North America), and 7(Europe). I_CpG varied significantly with CFR (P<0.0001) and these variations were highest with CFR of 4 but lowest with CFR of 7. Although the median I_CpG were close (CFR 1.2=0.408, CFR 2.1=0.4076, CFR 4=0.4072, CFR7=0.40755) and showed a downward trend with increasing CFR. CFR 7, however, had the lowest median I_CpG and showed the least variations. I_UpG varied significantly with CFR (P<0.0001) and the variations were highest with CFR of 4 and 1.2. CFR7, again, had the lowest median I_UpG and showed the least variations (Figure 3).

**Figure 3:**
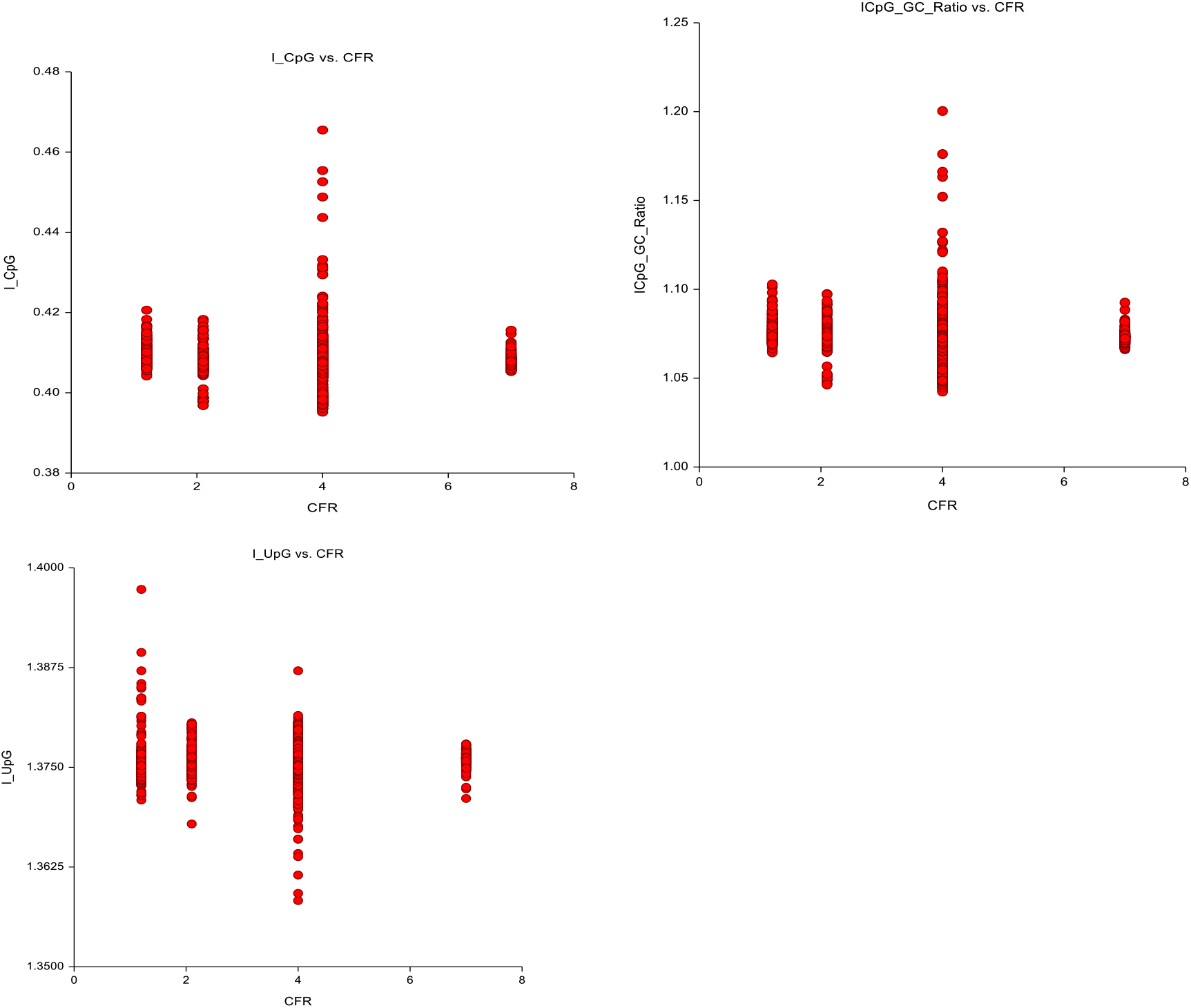
Variation of dinucleotide frequencies with crude case fatality rate (CFR) SARS-CoV-2 sequences from regions with CFR 4 show the highest I_CpG variability. This is not true with I_UpG.

### Comparison of variations of SARS-CoV-2 dinucleotide frequencies with patients’ status age and gender

There was significant difference in the I_CpG and GC% correlation between Asymptomatic (rs=0.675, 95% CI=0.560-0.764) and ICU/Deceased group (rs=0.778, 95% CI=0.719-0.825) (p<0.0001).

Regression of I_CpG on GC for Asymptomatic and ICU/Deceased appear divergent from a single point (Figure 4, 5), with ICU/Deceased group showing higher I_CpG for a given GC% (Slope-ICU/Deceased=46.14, R^2^= 0.72 and Slope Asy=7.17, R^2^= 0.58) (p<0.0001). I_UpG was significantly different between Asymptomatic and ICU/Deceased (P<0.0001) (Table 2). However, there was no significant difference between the medians of the I_CpG(p=0.25) of Asymptomatic and ICU/Deceased (Tables 2) (Figure 4,5).

**Figure 4.**
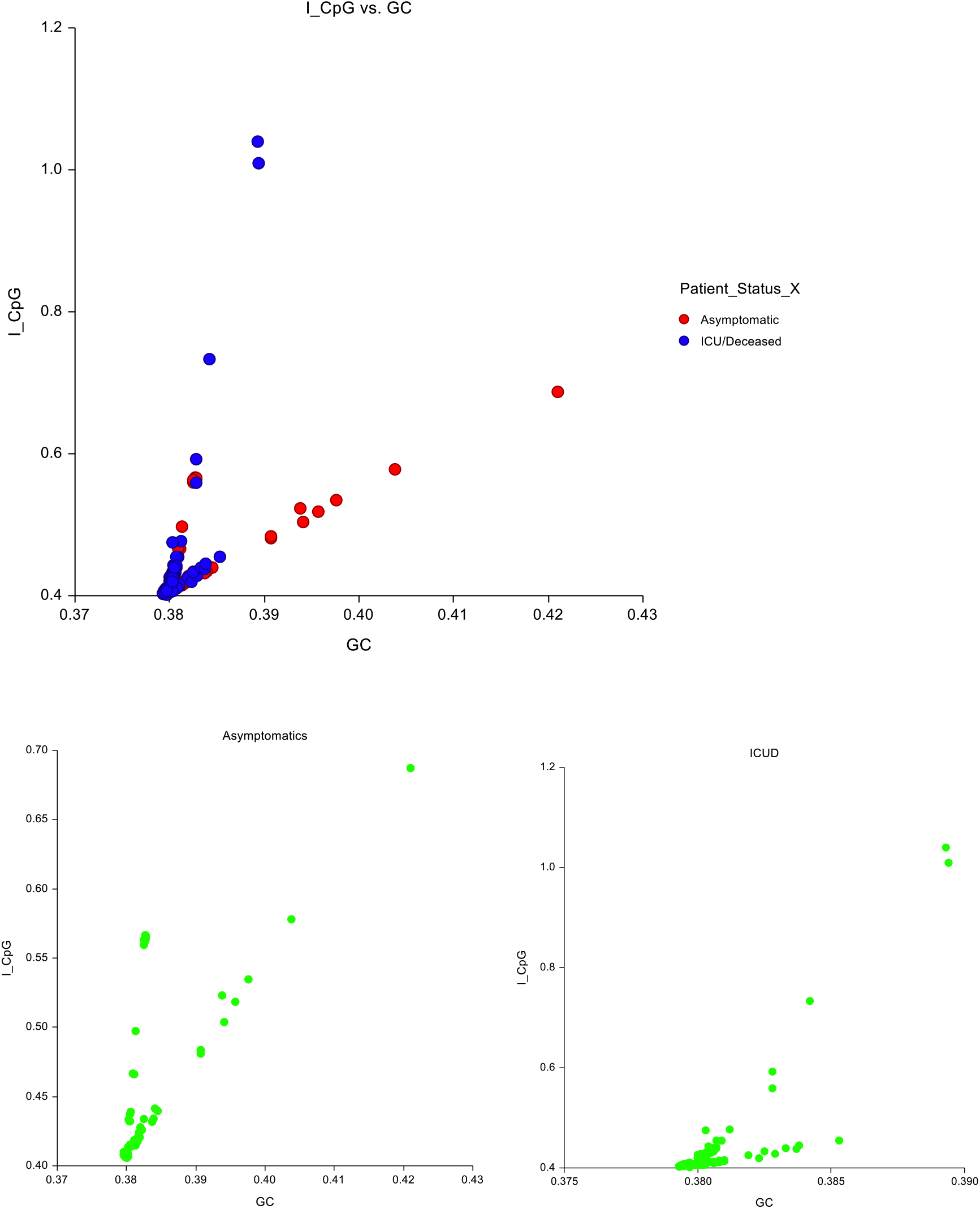
I_CpG Vs GC% distribution for Asymptomatic and ICU/Deceased COVID-19 patients. Asymptomatic: I_CPG=7.17GC-2.308, R^2^=0.58 and ICU/Deceased: I_CPG=46.14GC-17.12, R^2^=0.72. The rate of I_CpG variations with GC% of SARS-CoV-2 sequences from ICU/Deceased patients is almost 6.5 times that of sequences from asymptomatic patients suggesting highly competent ZAP activity in the later. The rate in asymptomatic is close to the background SARS-CoV-2 population rate of 7.3.

**Figure 5:**
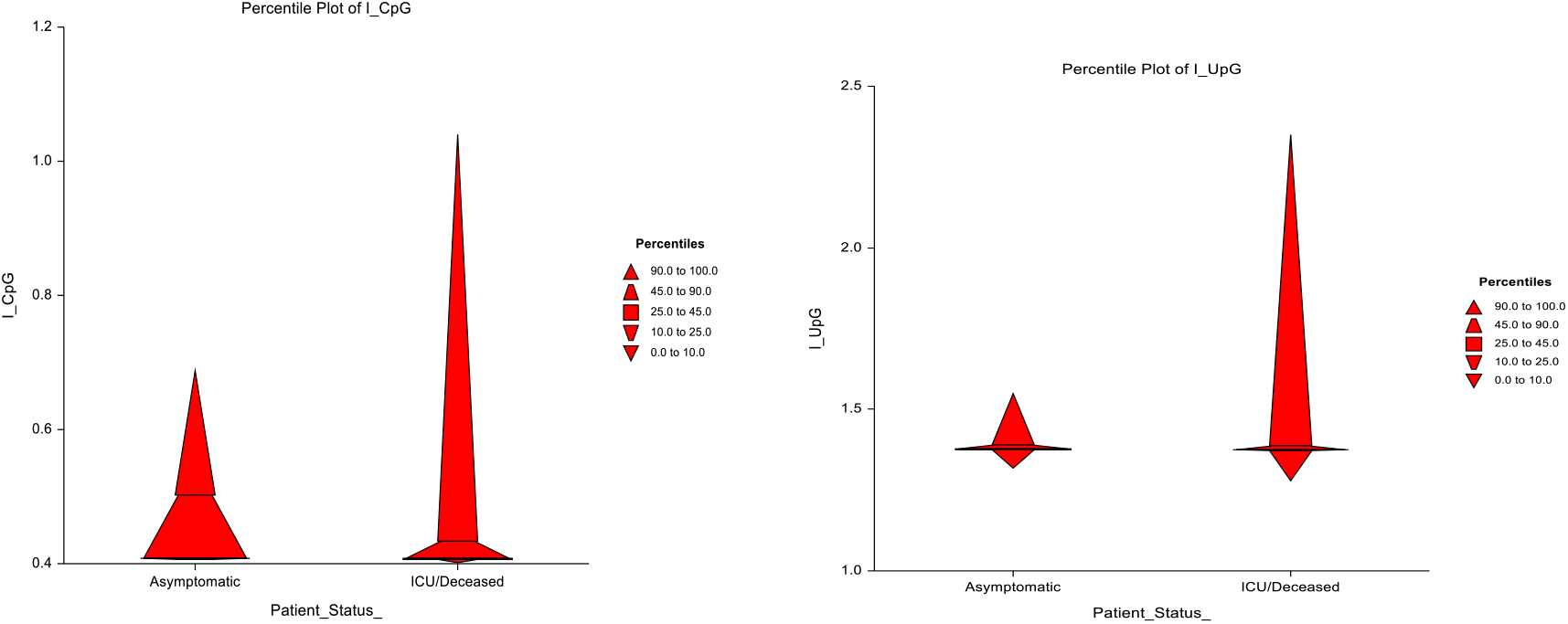
Percentile plot of I_CpG and I_UpG with patients’ status. The difference in I_CpG and I_UpG distribution between asymptomatic and ICU/Deceased patients is striking and suggests low ZAP competencies in the latter group.

**Table 2:** Variation of SARS-COV-2 dinucleotide frequencies with patients’ status N=349

| **I_CpG**<br>**by Patient Status** | **Minimum** | **1st Quartile** | **Median** | **3rd Quartile** | **Maximum** |
| --- | --- | --- | --- | --- | --- |
| Asymptomatic | 0.4058 | 0.40760 | 0.40790 | 0.42463 | 0.6872 |
| ICU/Deceased | 0.4013 | 0.40720 | 0.40825 | 0.42426 | 1.0398 |
| P=0.246 |  |  |  |  |  |
| **I_UpG**<br>**by Patient Status** | **Minimum** | **1st Quartile** | **Median** | **3rd Quartile** | **Maximum** |
| Asymptomatic | 1.3175 | 1.37550 | 1.37850 | 1.38140 | 1.5492 |
| ICU/Deceased | 1.2793 | 1.37460 | 1.37580 | 1.37813 | 2.3491 |
| P=0.0001 |  |  |  |  |  |

Of the 349 sequences with patient metadata from GISAID, the age of only 286 were available: Asymptomatic: 58 patients, Age: Mean=38.16 yrs., SD=18.75, Median=36yrs; ICU/Deceased: 228 patients, Age: Mean=65.48yrs, SD=16.37, Median=67 yrs. (Figure 6). I_CpG significantly varied with age-class with the elderly age class having the highest variations (I_CpG, p=0.0092) and middle-age group with the lowest variations (Figure 7). The age-group variations of I_UpG were not significant (p=0.45 and 0.058 respectively). There was no significant difference in SARS-CoV-2 dinucleotide distribution between genders; I_CpG(p=0.55) and I_UpG(p=0.49) (Figure 8).

**Figure 6:**
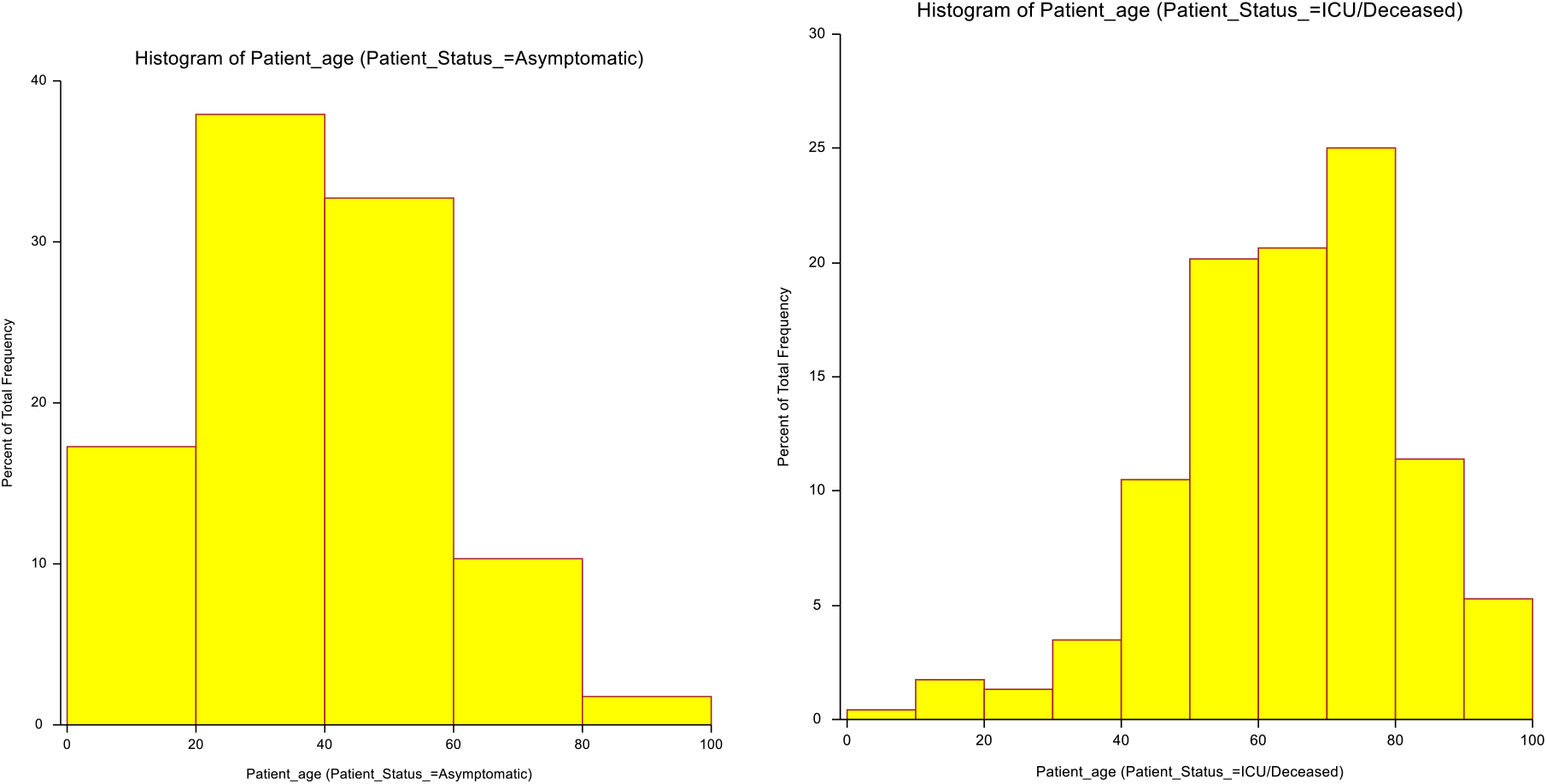
Age distribution by patients’ status. The age distribution pattern of asymptomatic and ICU/Deceased obtained from the metadata fits the pattern from previous studies, which reassures on the integrity of the data.

**Figure 7:**
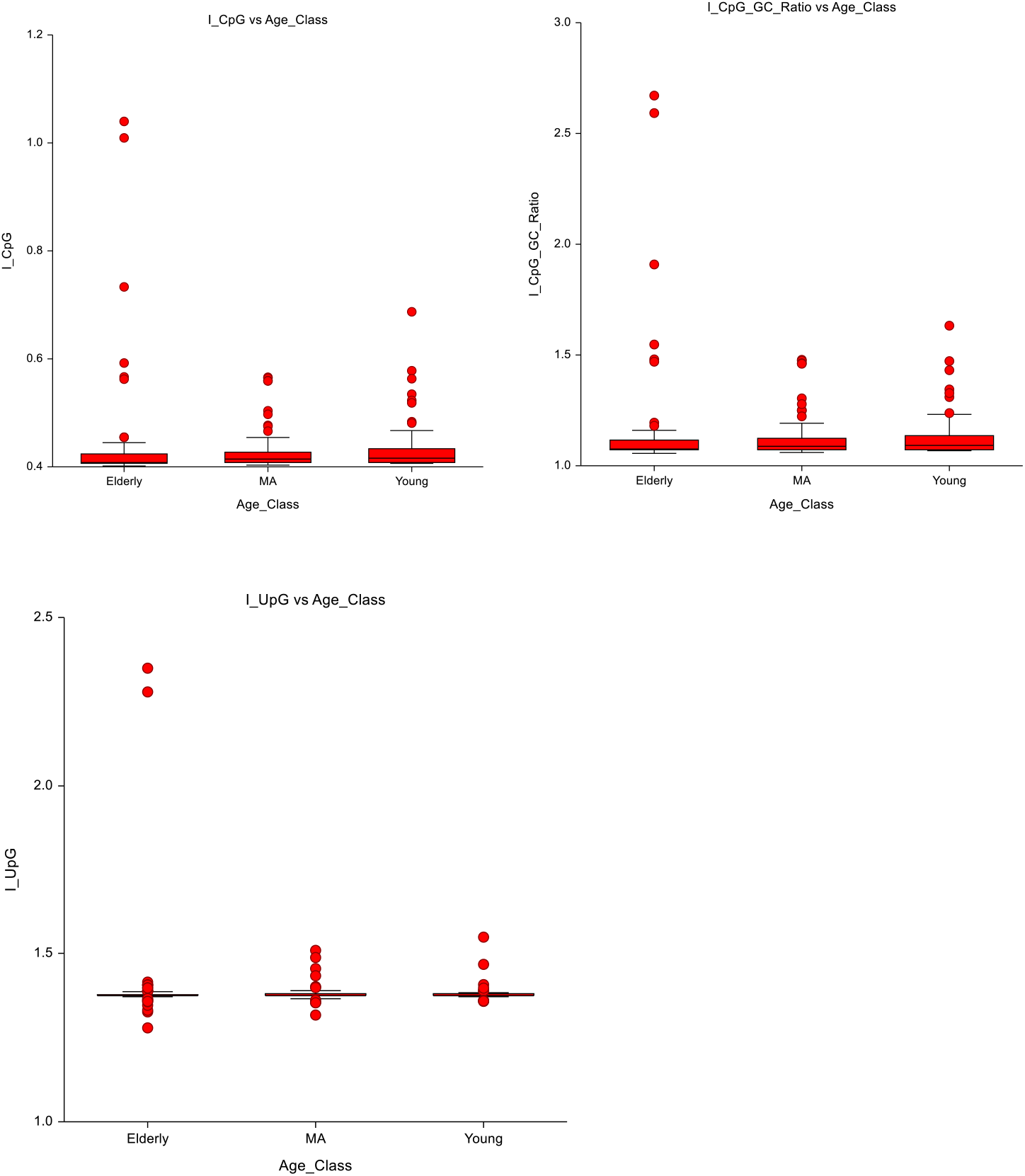
Variation dinucleotide frequencies with age-class. SARS-CoV-2 sequences from the elderly (>65yrs) show the highest I_CpG variability. The sequences from the middle age-class show the lowest variability. This is not true with I_UpG.

**Figure 8:**
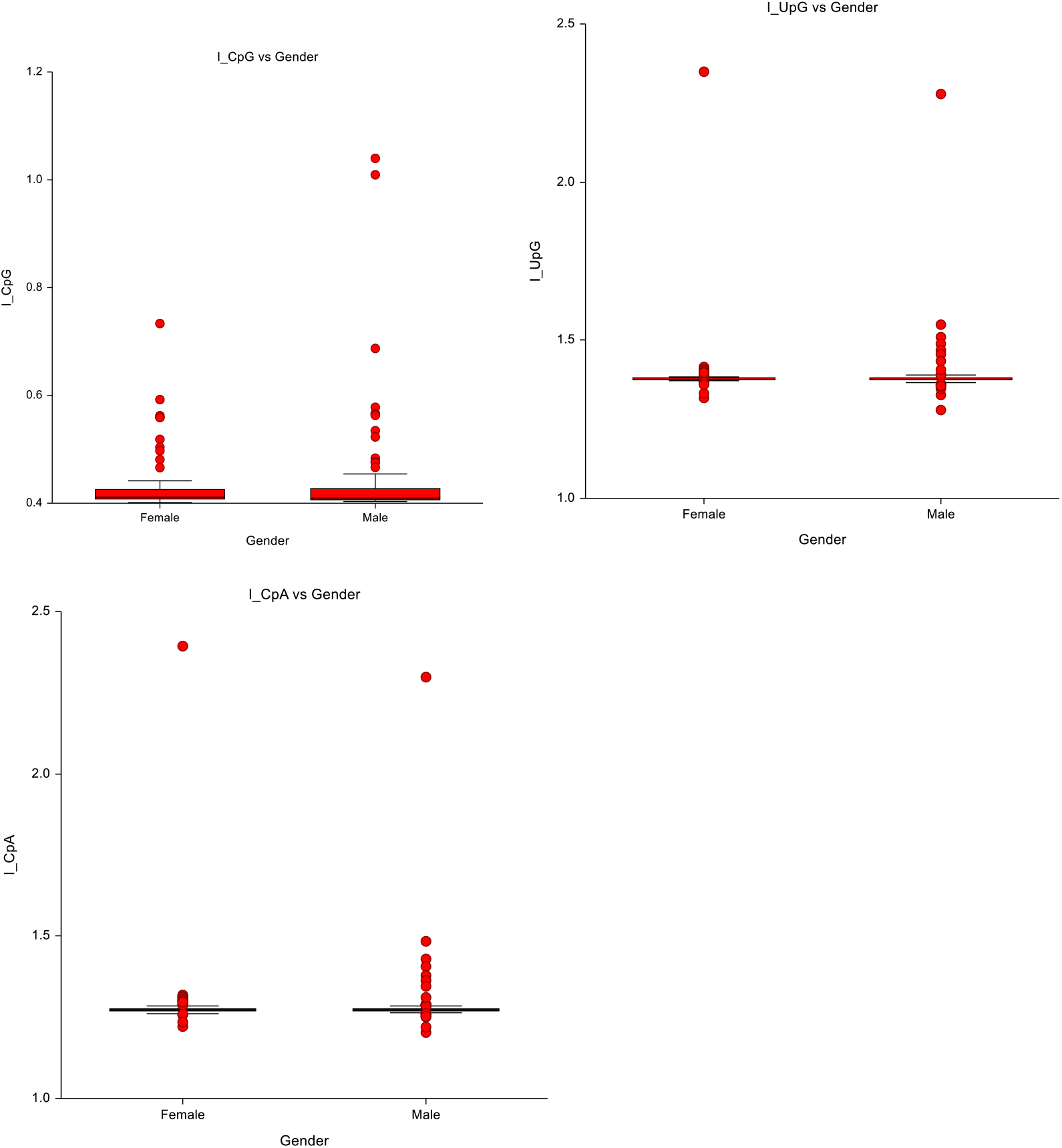
SARS_COV-2 dinucleotide distribution by patients’ gender. Though not statistically significant(P=0.55), the trend is towards higher I_CpG variability in the male gender, suggesting less competent ZAP activity in the male. This may partly explain the male preponderance of SARS-CoV-2 cases and the higher severity of COVID-19 in the male gender.

### Comparison of SARS-CoV-2 clades with patients’ status and with dinucleotide frequencies

SARS-CoV-2 Clade O is overrepresented in Asy (86/121) while SARS-CoV-2 Clades GR, GH, and G are overrepresented in ICU/Deceased (71/228,58/228, and 58/228 respectively) and the interaction of clade and patients’ status was significant(p<0.001) (Table 3). The I_CpG were significantly different between Clades (p<0.0006) with Clade GR having the highest I_CpG and variability compared to Clades GH, G, and O (Table 4). Furthermore, the variations of I_CpG with GC content was significantly different with clades(p<0.05) with Clade O having the lowest rate (Slope=8.56,R^2^=0.37) and highest rate with Clade GR (Slope=54.8, R^2^=0.88), Clade GH (Slope=22.8, R^2^=0.49) and Clade G (slope=16.3, R^2^=0.37).

**Table 3:** Contingency table of patients’ status versus SARS-COV-2 clades

|  | Patient Status |  |  |
| --- | --- | --- | --- |
| Clade | Asymptomatic | ICU/Deceased | Total |
| G | 12 | 58 | 70 |
| GH | 3 | 71 | 74 |
| GR | 12 | 58 | 70 |
| L | 0 | 6 | 6 |
| O | 86 | 21 | 107 |
| S | 8 | 12 | 20 |
| V | 0 | 2 | 2 |
| Total | 121 | 228 | 349 |
| Chi-Squared Tests |  |  |  |
|  | Value | df | p |
| X<sup>2</sup> | 152.786 | 6 | <0.001 |
| N | 349 |  |  |

**Table 4:**
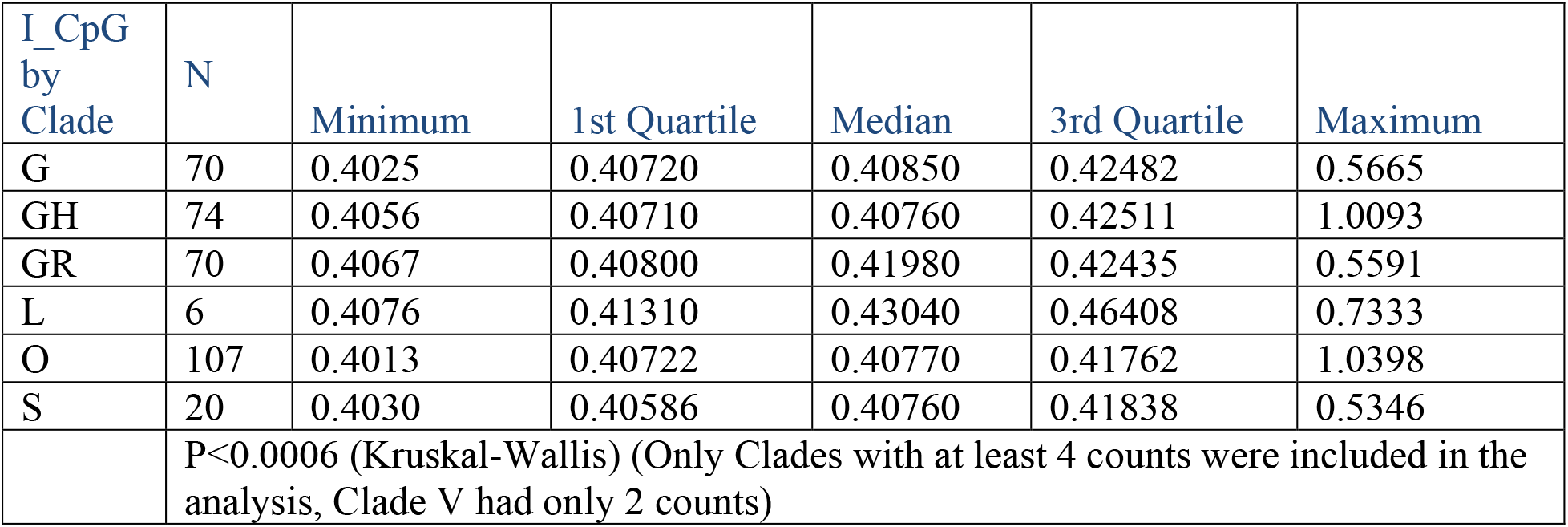
Variation of SARS-COV-2 dinucleotide frequencies with clades N=347

| I_CpG<br>by<br>Clade | N | Minimum | 1st Quartile | Median | 3rd Quartile | Maximum |
| --- | --- | --- | --- | --- | --- | --- |
| G | 70 | 0.4025 | 0.40720 | 0.40850 | 0.42482 | 0.5665 |
| GH | 74 | 0.4056 | 0.40710 | 0.40760 | 0.42511 | 1.0093 |
| GR | 70 | 0.4067 | 0.40800 | 0.41980 | 0.42435 | 0.5591 |
| L | 6 | 0.4076 | 0.41310 | 0.43040 | 0.46408 | 0.7333 |
| O | 107 | 0.4013 | 0.40722 | 0.40770 | 0.41762 | 1.0398 |
| S | 20 | 0.4030 | 0.40586 | 0.40760 | 0.41838 | 0.5346 |
| P<0.0006 (Kruskal-Wallis) (Only Clades with at least 4 counts were included in the analysis, Clade V had only 2 counts) |  |  |  |  |  |  |

### Sensitivity analysis

I_CpG-GC ratio had the same pattern of outcome (significant and non-significant) of comparisons as I_CpG. The results of the parametric analysis were the same pattern as non-parametric except that parametric analysis tends to overestimate test statistics (e.g. Spearman’s R^2^=0.357 for I_CpG to GC% correlation but Pearson’s R^2^=0.79).

## Discussion

Novel viruses in new hosts will more frequently encounter innate nonspecific host defense as the primary challenges[48], while the specific immune response lags[49]. The usual factors that dictate adequate host response include viral transmission load, viral infectivity, tissue tropism, host immune response, and viral susceptibility to therapeutics, among others. Though a large volume of knowledge has been accumulated on SARS-CoV-2 and COVID-19, there is still no clear explanation for skewed variations in morbidity and mortality as well as apparent discrepancies between ex-vivo viral susceptibility to candidate therapeutic and outcomes of clinical trials. Although a 382 nucleotide deletion has been associated with milder COVID-19, the prevalent D614G mutation, though increases transmission[9], does not appear related to disease morbidity[50]. However, mutation studies focus on protein-coding regions.

In this study, focused on whole genomic analysis, signatures of the differential activities of intracellular zinc finger antiviral protein (ZAP) appear to point to this intracellular protein as a key factor in the differential effect of SARS-CoV-2 on human hosts. It is possible to do this because ZAP activity leaves a signature in terms of SARS-CoV-2 genomic CpG index(I_CpG) such that in a highly restrictive environment (high ZAP activity) the I_CpG of SARS_CoV2 remains low and vice versa [16]. In this regard, this study has revealed that SARS-CoV-2 I_CpG has rapidly varied widely from the reference Wuhan virus and varied differentially between geographical regions consistent with widely different ZAP activities in humans. Differential human tissue and host ZAP activities have previously been shown in vitro studies of human cell lines[40], especially with regards to SARS-CoV-2 and target tissue viral production[16]. Interestingly African SARS-CoV-2 genome appears to have the lowest change in I_CpG to GC% correlation compared to the background SARS-CoV-2 population correlation (rs 0.357 vs rs 0.359, P>0.05) suggesting a highly restrictive ZAP environment. In support of this, I_CpG has the lowest absolute intersequence variation in Africa. Overall, the trend appears to be that areas with the highest ZAP activity have higher mortality except in Europe where regional SARS-CoV-2 intersequence I_CpG appears exceptionally low but has the highest crude case fatality. However, given that the I_CpG versus GC% correlation in Europe has widely varied from the SARS_CoV2 population correlation(rs=0.107 vs 0.359, P<0.05), one explanation is that though ZAP activity is low within the Europe population, the intra-host tissue variation in ZAP activity is low leading to low SARS-CoV-2 genomic I_CpG variability in Europe but overall, drift from reference(background) SARS-CoV-2 population I_CpG to GC% correlation.

To further test and give some validation to ZAP activity as a major factor in the outcome of SARS-CoV-2 infection, we evaluated the signature of ZAP activity in known risk factors whose data were available. In this regard, impressive I_CpG signatures of high ZAP activity and low ZAP activity were detected for Asymptomatic patients versus ICU/Deceased patients, respectively. Furthermore, Clade G(unsegregated), GR, and GH were overrepresented in ICU/Deceased patients and these clades also have I_CpG signature evidence of low ZAP activity. On the contrary, generic Clade O was overrepresented in asymptomatic patients and have I_CpG signature evidence of high ZAP activity. Interestingly, similar segregation is seen with age groups. Age group >65years, known to have a high risk of morbidity and mortality also have compelling I_CpG signature evidence of low ZAP activity. However, no significant differences in I_CpG signature of ZAP activity were seen between genders but the trend is towards higher variability in men, which is interesting because male gender appears to have up to 2.4 times higher risk of severe COVID-19 in some case series[51] though this was not supported by other studies [12].

UpG dinucleotide is also significantly differentially overexpressed in SARS-CoV-2 from different regions of the world. Furthermore, I_UpG also generally, but not consistently, segregated with regional case fatality and patents’ disease severity, age, and viral clade.

The public health implication of these findings is that while control of the spread of infection may depend on other factors, ZAP plays a major role in the outcome of SARS-CoV-2 infection, the disease phenotype, and probably treatment. In the first instance, ZAP activity levels are expected to be distributed within a given population in a Gaussian manner. Thus, in populations with overall high mean ZAP activity level, only 2.5% will be expected to be highly vulnerable while the remaining 97.5% would be susceptible to infection but with highly favourable outcomes mainly represented by mild illness, which may be missed as COVID-19, but leaves a high level of positive serology in the community. Nevertheless, flashes of severe illnesses may be seen in the 2.5% subset of such a population with lower levels of ZAP activity. Even, within countries, such flashes may be noted whenever in the country SARS-CoV-2 has recently been introduced. These appear to resemble the current SARS-CoV-2 scenario in Africa. However, in populations with an overall low mean ZAP activity level, more severe illness may be more common. Between these extremes, various scenarios may ensue all of which may be modified by other health system factors. Furthermore, it suggests that asymptomatic person probably paradoxically harbour potentially more virulent low CpG viruses which the asymptomatic is fitted to contain but the next person may not be.

The therapeutic implication is that candidate SARS-CoV-2 antiviral therapies may have a better chance of in-vivo success if they enhance ZAP activity and may have invitro versus invitro dissociation of efficacy if ZAP modulation is not one of its mechanism.

This may at least in part explain the increasing number of drugs showing impressive invitro activity against SARS-COV-2 but failing in trials even when started early (within 5 days of exposure) or even as pre-exposure prophylaxis. In this regard, it is interesting that strong interaction between Interferon and ZAP for its efficacy on COVID-19 has already been observed[16]. Although ZAP is an ISG product, ZAP also enhances the production of type 1 interferon[17]. Compromised ZAP competencies may, therefore, explain the low type 1 interferon response in severe COVID-19[52], the elderly[53], obesity[54], and type 2 diabetes[55]. ZAP compromise, through its modulation of Toll-like receptor[56], may also explain SARS-CoV-2 morbidity in hypertensives because these receptors are suggested to have a role in hypertensive morbidities[57]. Overall, these suggest that drugs (including interferon) that enhance ZAP activity, may favourably modulate the known poor outcome of SARS-CoV-2 infection in these groups of patients. Zinc sulphate has been found to have consistent invitro[58] and in vivo effect against SARS-COV-2[59] and amongst many possible pathways for the effect of elemental zinc, it is interesting that it also enhances the production of interferon[59]. Dexamethasone is strongly anti-inflammatory and inhibits interferon[60], thus reducing ZAP activity. Dexamethasone may, therefore, may be predicted to be harmful in mild COVID-19, when viral replication appears to drive morbidity and beneficial in severe COVID-19 when hyper-inflammation and cytokine storm appears to drive morbidity. This probably explains the finding of the effect of dexamethasone in the ‘Recovery trial’[37]. The funding further suggests that going forwards, arms of clinical trials may include a combination of interferon or its inducers, anti-viral, and zinc for mild to moderate cases and the addition of anti-inflammatory agents for severe cases.

Given the findings in this study, it is tempting to propose that communities with higher COVID-19 mortalities and those with lower mortalities have been pointed in the respective direction by the innate ZAP activity status while health system effectiveness may change or worsen the directions. It may also be suggested that in patients with lower ZAP competencies, SARS-CoV-2 infection in sanctuary sites, like CNS, may represent a slow smouldering disease, that may be better described as subclinical instead of asymptomatic, with ominous possible long term consequences. Given such scenarios, prevention, vaccine, prophylaxis, and therapeutics may need to be prioritized in that order.

Overall, our study suggests that innate intracellular antiviral activities may be a major determinant of the outcome of SARS-CoV-2 infection. This consideration appears to explain many puzzling observations regarding SARS-CoV-2 and COVID-19. In this regard, it may be important to examine how ZAP levels, forms, distribution, and activities are influenced by factors such as genetic polymorphism, race, co-morbidities, and aging to inform net ZAP activity as a contributing determinant of severity of Covid-19 phenotype.

The limitations of this study are that database bias, group size bias and over-interpretation may result in spurious findings. Nonetheless, this study provides an additional explanation to the confounding observations in a disease of certain uncertainties: COVID-19. Prospective studies will be required to provide further validation of our findings.

## Data Availability

All data for the amnuscript are available at the URL below

https://drive.google.com/drive/folders/1q1jVVp9u2G-AT2MZYRsYD8fDqN8HIb3R?usp=sharing

## Acknowledgments

We acknowledge the following for reviewing the manuscript: Drs. Peters Wale Oladosu, Mohammad Asara Abdullahi, Clement Meseko and Kelly Osezele Elimian, of the Nigeria COVID-19 Research Consortium.

## Author contributions

Shaibu Oricha Bello conceived and developed the idea, ran the sequence analysis, and drafted the initial manuscript. Ehimario Igumbor, Yusuf Deeni, Chinwe Lucia Ochu, and Popoola Mustapha Ayodele reviewed the manuscripts and expanded on the arguments. Ehimario Igumbor re-examined the ethics of the study. All authors contributed to the final manuscript.

## Competing Interests

The authors declared no competing interests.

## Materials & Correspondence

Address all correspondence and materials requests to Shaibu Oricha Bello, Department of Pharmacology & Therapeutics, Usmanu Danfodiyo University, *UDUTH-Campus, 1 GarbaNadamaRoad, Sokoto. Nigeria.*

## Notes

### Competing Interest Statement

The authors have declared no competing interest.

### Funding Statement

No external funding was received for this study

### Author Declarations

Ethical exemption was obtained from UDUS ethical board

